# Marker or mechanism? Medical imaging utilisation and population life expectancy in 38 OECD countries, 2000–2024 The RAD-LIFE study

**DOI:** 10.64898/2026.09.02.26362082

**Authors:** Rafał Obuchowicz

## Abstract

**Background:** Medical imaging is central to modern diagnosis and treatment, yet its contribution to population-level survival has rarely been quantified, and the few existing estimates have been interpreted as evidence that imaging directly extends life. Whether national imaging utilisation reflects a distinct clinical mechanism or a broader process of health-system development has not been formally tested.

**Methods:** Longitudinal ecological panel study of 38 OECD countries, 2000–2024 (950 possible country-years). CT and MRI availability and utilisation were linked to life expectancy at birth, treatable and preventable mortality, and seven cause-specific mortality outcomes. Two-way fixed-effects models with country and year fixed effects and country-clustered standard errors were adjusted sequentially for log GDP per capita, log health expenditure per capita, population aged ≥65 years, physician density and smoking prevalence. Three falsification families were specified: lead-exposure (placebo) models regressing current life expectancy on future imaging utilisation; negative control exposures with no diagnostic-imaging pathway to survival; and wild cluster bootstrap inference.

**Results:** In the principal adjusted model, CT utilisation at a three-year lag was associated with higher life expectancy (β = 0.85 years per 100 additional examinations per 1,000 inhabitants, 95% CI 0.16 to 1.54; cluster-robust p = 0.015; bootstrap p = 0.034; n = 339, 29 countries). Scanner availability showed weaker, generally non-significant associations. All three falsification analyses failed to support a modality-specific effect. Future CT utilisation predicted current life expectancy as strongly as past utilisation (three-year lead β = 0.86, 95% CI 0.15 to 1.57, p = 0.017), consistent with high within-country autocorrelation of the exposure (r = 0.64 at three years). Radiotherapy equipment density, a therapeutic rather than diagnostic technology, showed an equal and more precise association (β = 0.85 per 10 units per million, 95% CI 0.32 to 1.38, p = 0.0017), as did PET scanner density and health expenditure, whereas physician density showed none. Associations were stronger for preventable (β = −43.3 per 100,000, p = 0.0002) than treatable mortality (β = −13.5, p = 0.19), and all were attenuated to the null under country-specific linear trends.

**Conclusions:** National CT utilisation is a robust ecological correlate of life expectancy but does not behave as a causal determinant under falsification. It is best understood as a marker of health-system capital development. Claims that imaging adds years of life are not supportable on ecological evidence of this kind.

## 1. Introduction

Medical imaging has become one of the central infrastructures of modern healthcare. Computed tomography (CT), magnetic resonance imaging (MRI), ultrasonography, nuclear medicine and image-guided procedures support diagnosis, treatment selection, disease monitoring, screening, emergency care, oncology, cardiovascular medicine and trauma management. In many clinical pathways imaging is no longer an adjunct but a prerequisite for timely and accurate decision-making.

Despite this central role, the population-level contribution of radiology to health outcomes remains difficult to quantify. Most evaluations of imaging focus on diagnostic accuracy, workflow efficiency, time to diagnosis, downstream clinical management or cost-effectiveness. These endpoints are important, but they do not answer a broader question: does greater access to and utilisation of imaging translate into measurable improvements in population survival?

The small existing literature on this question has generally answered in the affirmative. Analyses of United States state-level panel data reported that longevity increased more rapidly in states with faster growth in the share of advanced diagnostic imaging procedures, with imaging attributed roughly 0.6–0.7 years of a 2.4-year gain in life expectancy between 1991 and 2004 [1,2]. Comparable analyses have been reported for other national settings [3], and regional ecological studies have reported associations between the density of specific imaging services and cause-specific mortality [4]. These results have been widely cited in advocacy for imaging capacity. The design family they belong to has attracted methodological criticism, principally that the underlying regressions are vulnerable to misspecification and to correlated secular trends [5], but that criticism has not been resolved empirically. In particular, these analyses have not been subjected to the falsification tests that are now standard in observational health-economics research, and the counter-hypothesis — that imaging utilisation is a proxy for the general developmental state of a healthcare system — has not been formally evaluated.

That counter-hypothesis is not merely theoretical. National income has long been recognised as a dominant correlate of population longevity [6], and cross-country panel analyses of health expenditure, workforce and lifestyle determinants of life expectancy consistently show strong collinearity among health-system inputs [7,8]. Countries with greater imaging capacity are also wealthier, spend more on healthcare, employ more physicians and have better organised health systems. Simple cross-sectional associations between scanner density and life expectancy are therefore highly susceptible to confounding, and longitudinal within-country analysis is a minimum requirement. But even within-country analysis is vulnerable when the exposure evolves as a smooth national trend, because the exposure then carries little information beyond the passage of time within that country.

A second, more tractable distinction concerns technology availability versus technology use. National imaging capacity and imaging volume are reported as separate indicators and vary markedly and non-proportionally between countries [33]: some healthcare systems possess large numbers of scanners but relatively low examination volumes, whereas others integrate imaging more intensively into routine care. Availability and utilisation can therefore be separated empirically, and their divergence is informative about whether infrastructure alone carries population-level meaning.

The present study was designed to evaluate the longitudinal association between national imaging capacity and utilisation and population survival across OECD countries, and — critically — to test whether any such association behaves as a modality-specific effect or as a marker of broader health-system capacity. We specified three falsification strategies for this purpose: lead-exposure (placebo) models, in which current survival is regressed on future imaging utilisation; negative control exposures, comprising health-system resources with no diagnostic-imaging pathway to survival; and cause-specific analyses testing whether associations concentrate in conditions where imaging is clinically decisive.

## 2. Materials and methods

### 2.1 Study design

We conducted a longitudinal ecological panel study of the association between national medical imaging capacity and utilisation and population-level survival across member countries of the Organisation for Economic Co-operation and Development (OECD). The unit of analysis was the country-year. All 38 OECD member countries were eligible; annual observations were collected for 2000–2024 where available, giving 950 possible country-years. Because reporting completeness differed across countries, years and indicators, primary analyses used an unbalanced panel, retaining all country-years for which the required variables were simultaneously observed rather than restricting analysis to countries with complete series. The study used exclusively publicly available aggregated national data and did not involve individual patient data.

### 2.2 Data sources

Imaging variables and population health outcomes were obtained from OECD Health Statistics [9]: imaging capacity from the Medical Technology Availability dataset, imaging utilisation from the Diagnostic Exams dataset, and outcomes from the Life Expectancy, Avoidable Mortality and Causes of Mortality datasets. Time-varying socioeconomic, demographic and health-system covariates were obtained from the World Bank World Development Indicators [10]: GDP per capita (current US), *current health expenditure per capita(currentUS)* physicians per 1,000 population, population aged ≥65 years as a percentage of the total, and prevalence of current tobacco use among adults. Sources were harmonised by ISO3 country code and calendar year and merged into a single country-year panel.

### 2.3 Imaging exposures

Imaging capacity was represented by CT scanners and MRI units per 1,000,000 inhabitants; imaging utilisation by CT and MRI examinations per 1,000 inhabitants. Total healthcare-provider values were used. For interpretability, capacity variables were scaled per 10 additional units per million inhabitants and utilisation variables per 100 additional examinations per 1,000 inhabitants. Because effects of increased diagnostic capacity may occur with delay, exposures were evaluated contemporaneously and with 1-, 3- and 5-year lags.

### 2.4 Population outcomes

The principal outcome was life expectancy at birth (years, total population). Secondary outcomes were treatable and preventable mortality (age-standardised deaths per 100,000). Both are defined by the joint OECD/Eurostat lists of avoidable causes of death [11], which build on earlier work on mortality amenable to healthcare [12,13] and restrict avoidable deaths to those occurring below 75 years of age. Treatable mortality comprises causes largely avoidable through timely and effective healthcare, including secondary prevention; preventable mortality comprises causes largely avoidable through public health and primary prevention. The age restriction means these outcomes capture premature mortality only, whereas life expectancy at birth reflects mortality at all ages. Exploratory cause-specific outcomes were cerebrovascular diseases, ischaemic heart diseases, neoplasms, malignant neoplasms of trachea/bronchus/lung, transport accidents, accidental falls and accidents overall, all as age-standardised rates per 100,000.

### 2.5 Covariates

The core adjustment set comprised log GDP per capita, log health expenditure per capita and the proportion of population aged ≥65 years. GDP and health expenditure were natural-log transformed because both were strongly right-skewed and represent multiplicative differences in national resources. An extended model added physicians per 1,000 population. Smoking prevalence was added in a restricted sensitivity analysis. No obesity variable was included because consistent country-year coverage was unavailable.

### 2.6 Missing data

Data availability was assessed separately for each variable. No values were imputed in the primary analyses; models used a complete-case unbalanced panel at model level. Because smoking prevalence was reported intermittently, smoking-adjusted analyses were complete-case without temporal interpolation. This has a consequence that we quantify explicitly rather than assume away: adding smoking changes the analytical sample as well as the adjustment set, and the two effects are separable (Section 3.4).

### 2.7 Fixed-effects panel models

Principal analyses used two-way fixed-effects regression with country and calendar-year fixed effects:

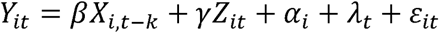

Where *Y_it_* is the outcome for country i in year *t,X_i,t-k_* the imaging exposure at lag k, *Z_it_* time-varying covariates, *α_i_* country fixed effects and *λ_t_* year fixed effects. Country fixed effects absorb all approximately time-invariant national characteristics; year fixed effects absorb global secular change. Standard errors were clustered at country level.

Two-way fixed-effects regression with a continuously distributed, non-staggered exposure imposes linearity and homogeneity of the exposure–outcome relationship, and recent econometric work has shown that such estimators can be difficult to interpret when treatment effects are heterogeneous across units or over time [14–16]. We therefore treat the estimated coefficients as linear approximations to an average within-country association rather than as structural parameters, and we rely on falsification analyses rather than on the fixed-effects specification alone to assess whether a causal reading is tenable.

### 2.8 Sequential adjustment

Models were built sequentially: Model 1, country and year fixed effects; Model 2, plus log GDP per capita, log health expenditure per capita and population ≥65 years; Model 3, plus physician density; Model 4, plus smoking prevalence (complete-case).

### 2.9 Lead-exposure (placebo) analyses

To test whether lagged associations reflect temporal precedence rather than shared secular trends, we estimated lead-exposure models in which life expectancy in year t was regressed on CT utilisation in years t+1, t+3 and t+5, using the extended specification. A genuine lagged effect cannot operate backwards in time; a lead association of comparable magnitude therefore indicates that the lagged estimate is not identifying a directional effect, an argument that parallels the standard use of pre-trend and placebo tests in panel-data analysis [24]. To characterise the information content of the exposure we also computed the within-country autocorrelation function of CT utilisation after removal of country and year means.

### 2.10 Negative control exposures

To distinguish a modality-specific contribution of CT from a generic association between health-system capacity and longevity, the extended specification was re-estimated with CT utilisation replaced by exposures that share the determinants of national health-system investment but lack a diagnostic-imaging pathway to population survival: radiotherapy equipment per million inhabitants (a therapeutic technology), gamma cameras per million, mammographs per million, PET scanners per million, physicians per 1,000 population, and log health expenditure per capita. If the association observed for CT reflects diagnostic imaging specifically, these controls should be null; if it reflects health-system capital development, they should behave similarly [17,18].

### 2.11 Negative control outcomes

Cause-specific analyses served a parallel falsification role. Associations were expected to concentrate in conditions for which imaging is clinically decisive — for example low-dose CT screening in lung cancer [25,26] and imaging-based patient selection for endovascular thrombectomy in acute ischaemic stroke [27] — and to be absent for outcomes with no diagnostic-imaging pathway. Because 112 model comparisons were evaluated, Benjamini– Hochberg false-discovery-rate correction was applied across this family.

### 2.12 First-difference and country-trend models

First-difference models related annual within-country changes in exposure to annual changes in outcome, with year fixed effects and country-clustered standard errors. Separately, models incorporating country-specific linear time trends in addition to country and year fixed effects were estimated, allowing each country its own long-term trajectory. These are stringent specifications that may also absorb genuine slow-acting effects and are therefore reported as robustness analyses.

### 2.13 Within-country descriptive analysis

Imaging and outcome variables were demeaned within country and year, and correlations between the resulting deviations examined, to establish whether observed associations were driven by within-country change rather than static between-country differences.

### 2.14 Restricted secondary analysis

Following the exploratory stage, six biologically motivated associations were evaluated with the full adjustment set including smoking, with Holm correction across the six comparisons. Because these hypotheses were selected after inspection of the exploratory results, this stage is designated a restricted hypothesis-driven validation analysis rather than an independently pre-registered confirmatory analysis.

### 2.15 Inference

Effects are reported as regression coefficients with 95% confidence intervals and two-sided p values. Because the number of clusters is modest (29 countries for utilisation models), cluster-robust standard errors may be anti-conservative; inference for the principal models was therefore repeated using a wild cluster bootstrap with Rademacher weights and the null imposed (B = 1,999) [19,20]. A two-sided p < 0.05 was considered significant for individual analyses; multiplicity-adjusted p or q values were used for model families. Given the ecological design, all coefficients are interpreted as associations rather than causal effects.

### 2.16 Software

Panel construction, descriptive statistics, correlation analyses, regression modelling and bootstrap inference were performed in Python using ordinary least-squares estimation with cluster-robust covariance. Analysis code and the constructed country-year panel are available as supplementary material.

## 3. Results

### 3.1 Dataset composition and completeness

The analytical dataset comprised 38 OECD countries over 2000–2024 (950 possible country-years). Completeness varied markedly across domains (Table 1). Life expectancy was available for 947 country-years (99.7%) and treatable and preventable mortality for 868 (91.4%) each. Imaging capacity was available for 736 country-years (77.5%, 37 countries) for CT scanners and 694 (73.1%, 36 countries) for MRI units. Utilisation data were substantially sparser: CT examination rates for 493 country-years (51.9%) and MRI examination rates for 475 (50.0%), each across 29 countries. Among covariates, GDP per capita and population ≥65 years were complete, health expenditure was available for 934 country-years (98.3%) and physician density for 835 (87.9%). Smoking prevalence was available for only 342 country-years (36.0%). No country had complete information for all principal variables across the full period; a fully observed balanced subset comprised 16 countries over 2015–2023.

**Table 1.** Data sources, variable definitions and completeness of the RAD-LIFE dataset

| Domain | Variable | Unit / definition | Countries | Observed | Possible | Complete |
| --- | --- | --- | --- | --- | --- | --- |
| Imaging exposure | CT scanner availability | Units per 1,000,000 inhabitants | 37 | 736 | 950 | 77.5% |
| Imaging exposure | MRI scanner availability | Units per 1,000,000 inhabitants | 36 | 694 | 950 | 73.1% |
| Imaging exposure | CT utilisation | Examinations per 1,000 inhabitants | 29 | 493 | 950 | 51.9% |
| Imaging exposure | MRI utilisation | Examinations per 1,000 inhabitants | 29 | 475 | 950 | 50.0% |
| Negative control exposure | Radiotherapy equipment | Units per 1,000,000 inhabitants | 33 | 579 | 950 | 60.9% |
| Negative control exposure | Mammographs | Units per 1,000,000 inhabitants | 34 | 587 | 950 | 61.8% |
| Negative control exposure | Gamma cameras | Units per 1,000,000 inhabitants | 34 | 578 | 950 | 60.8% |
| Negative control exposure | PET scanners | Units per 1,000,000 inhabitants | 35 | 628 | 950 | 66.1% |
| Outcome | Life expectancy at birth | Years, total population, age 0 | 38 | 947 | 950 | 99.7% |
| Outcome | Treatable mortality | Age-standardised deaths per 100,000 | 38 | 868 | 950 | 91.4% |
| Outcome | Preventable mortality | Age-standardised deaths per 100,000 | 38 | 868 | 950 | 91.4% |
| Core confounder | GDP per capita | Current US, *log – transformed*\|38\|950\|950\|1, log-transformed | 38 | 934 | 950 | 98.3% |
| Core confounder | Physician density | Physicians per 1,000 population | 38 | 835 | 950 | 87.9% |
| Core confounder | Population aged ≥65 years | % of total population | 38 | 950 | 950 | 100.0% |
| Sensitivity confounder | Current tobacco use | % of adults, complete-case only | 38 | 342 | 950 | 36.0% |

*Study frame: 38 OECD member countries, 2000–2024. The primary analysis used an unbalanced panel. Smoking prevalence was analysed without interpolation.*

### 3.2 Evolution of imaging utilisation

Imaging utilisation rose substantially and heterogeneously across the study period (Figure 1). Median CT utilisation across reporting OECD countries approximately doubled between 2000 and 2024, with wide and persistent between-country dispersion; MRI utilisation followed a similar but flatter trajectory. Within-country series were smooth: after removal of country and year means, the autocorrelation of CT utilisation was 0.86 at one year, 0.64 at three years, 0.45 at five years and 0.35 at six years.

**Figure 1.**
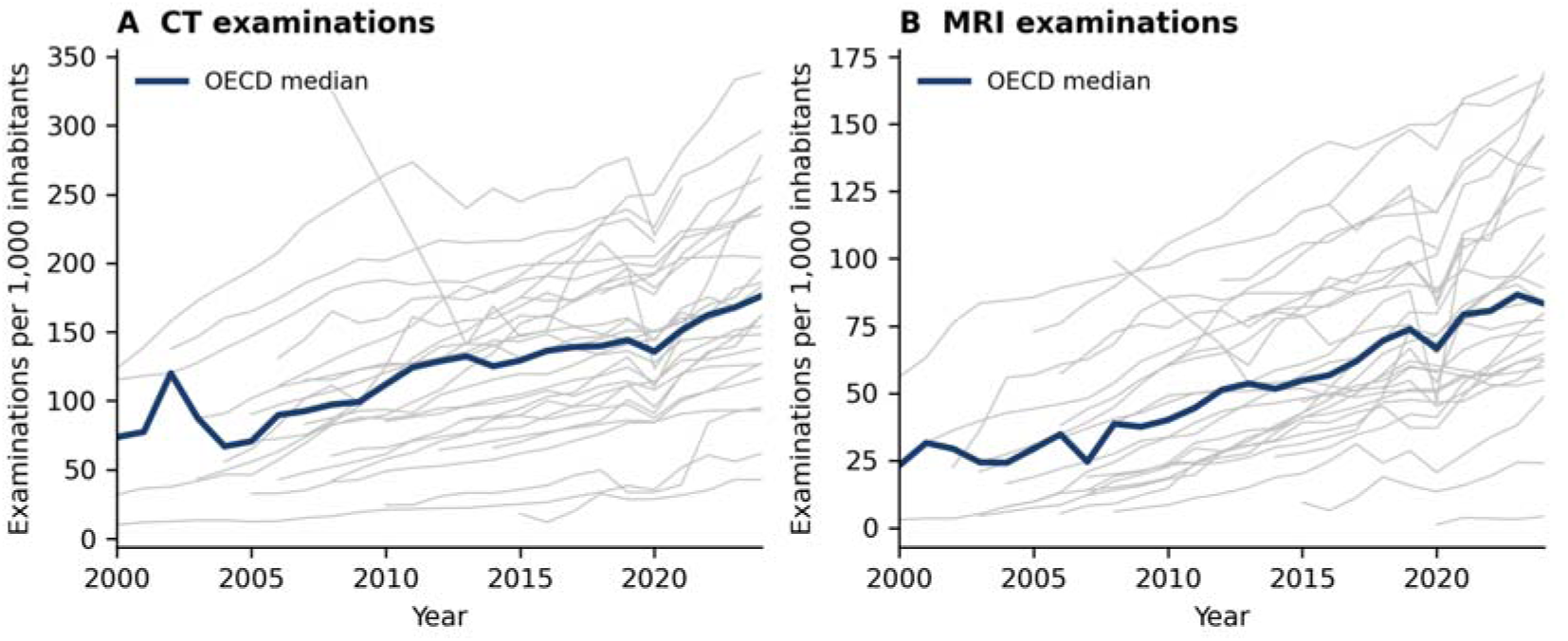
Evolution of imaging utilisation across OECD countries, 2000–2024. (A) CT examinations per 1,000 inhabitants; (B) MRI examinations per 1,000 inhabitants. Grey lines are individual countries; the dark line is the annual OECD median across reporting countries.

### 3.3 Within-country association and sequentially adjusted models

Within-country deviations in CT utilisation were positively associated with within-country deviations in life expectancy (Figure 2), confirming that the relationship is not solely a static between-country contrast.

**Figure 2.**
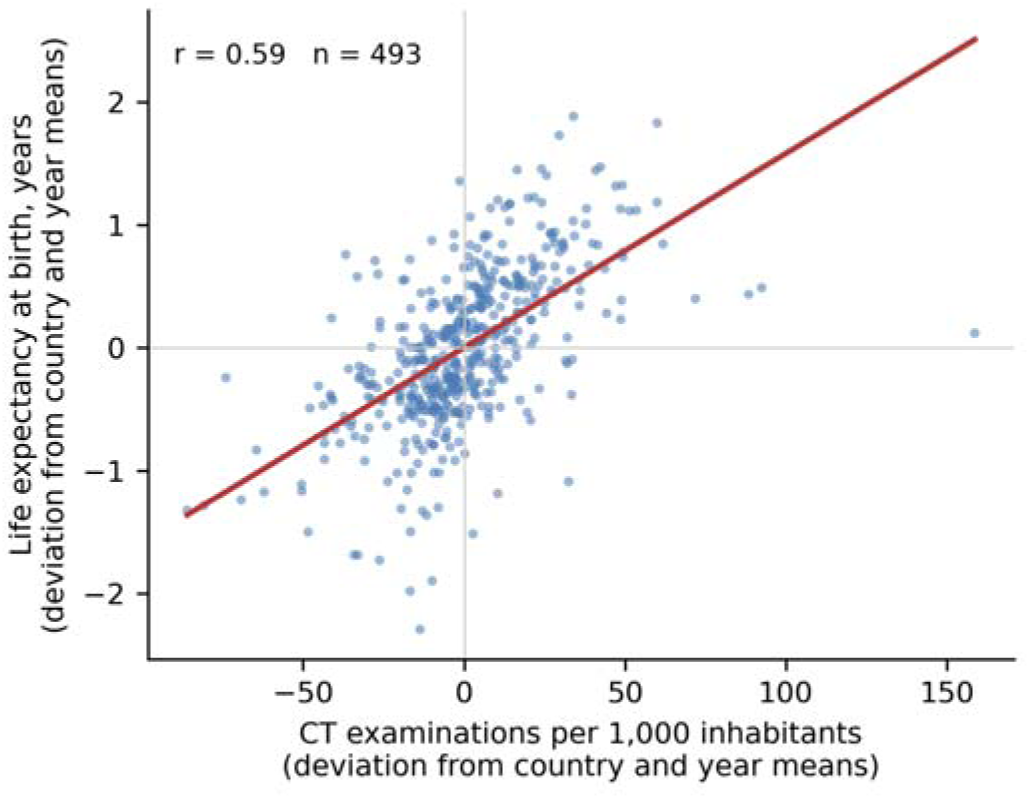
Within-country relationship between CT utilisation and life expectancy. Each point is one country-year, expressed as a deviation from that country’s mean and from the corresponding year mean. The line is the ordinary least-squares fit.

In pooled country-year analyses, imaging capacity and utilisation were generally associated with more favourable outcomes; MRI scanner availability correlated inversely with treatable mortality (Pearson r = −0.50, Spearman ρ = −0.60, both p < 0.001) and CT utilisation with treatable (r = −0.23, p < 0.001) and preventable mortality (r = −0.13, p = 0.004). These pooled correlations are descriptive only.

After introducing country and year fixed effects, associations between scanner availability and outcomes attenuated markedly. For CT scanner availability, an increase of 10 scanners per million inhabitants was not significantly associated with life expectancy at any lag (lag 0: β = 0.10 years, 95% CI −0.21 to 0.41, p = 0.53; lag 3: β = 0.17, p = 0.39; lag 5: β = 0.22, p = 0.31), nor with treatable or preventable mortality.

Imaging utilisation retained a stronger association with survival (Table 2). At a three-year lag, an increase of 100 CT examinations per 1,000 inhabitants was associated with 0.81 additional years of life expectancy under country and year fixed effects (95% CI 0.29 to 1.32, p = 0.0020), 0.86 years after socioeconomic adjustment (95% CI 0.22 to 1.50, p = 0.0086) and 0.85 years after further adjustment for physician density (95% CI 0.16 to 1.54, p = 0.0154). Under wild cluster bootstrap inference the corresponding p value for the extended model was 0.034. MRI utilisation attenuated with adjustment and remained non-significant throughout (extended model β = 0.23 years, 95% CI −1.14 to 1.60, p = 0.742).

**Table 2.**
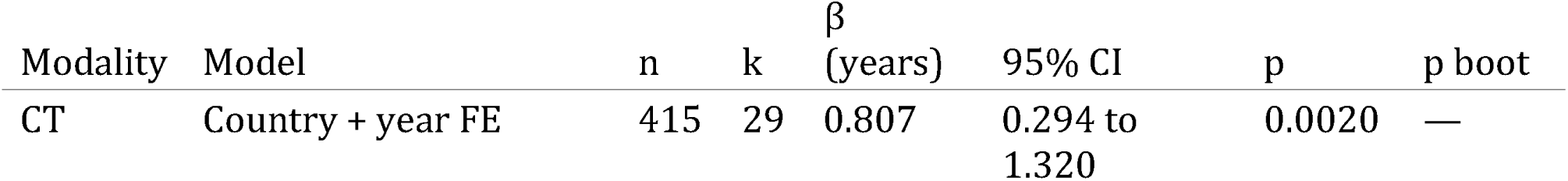
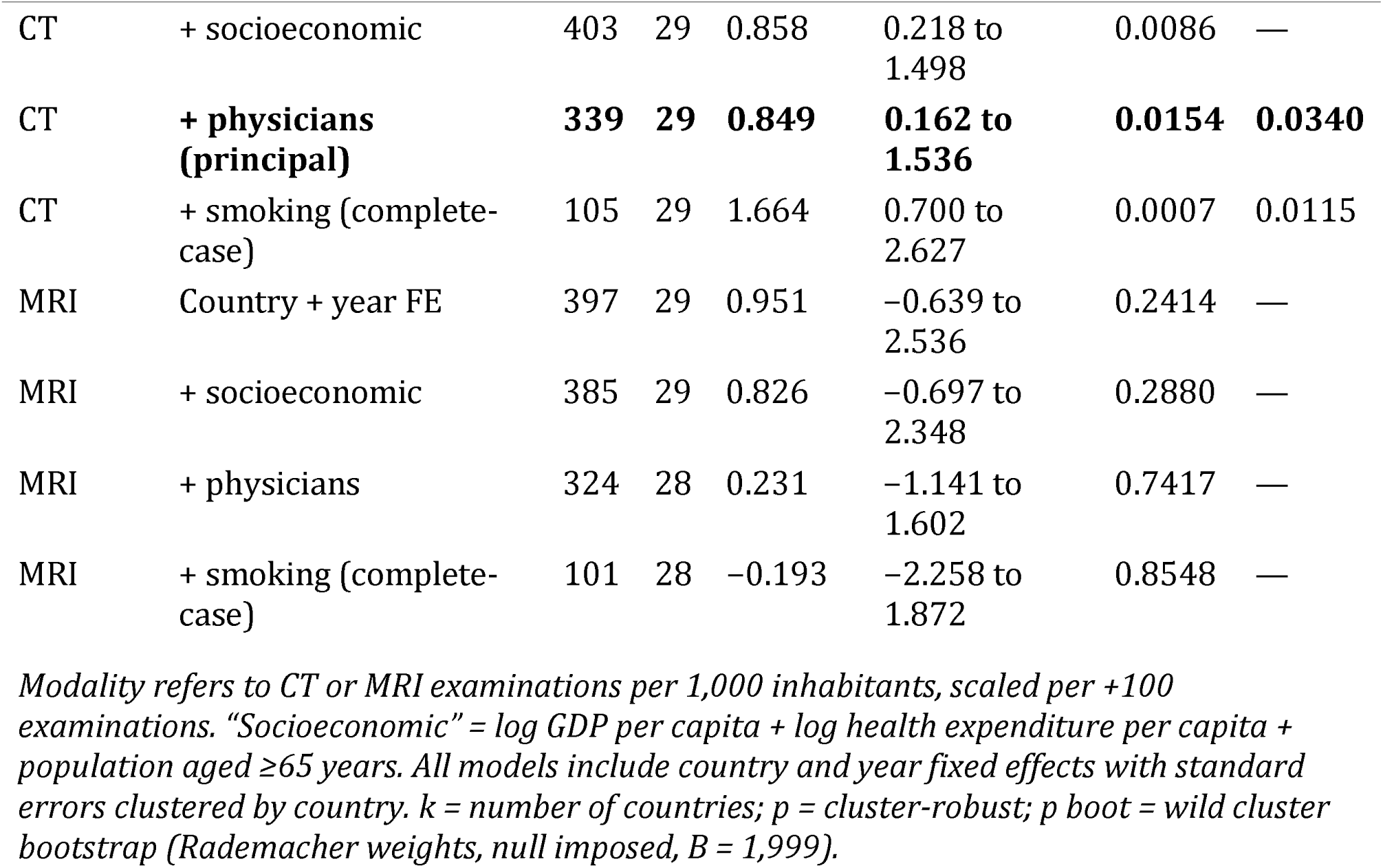
Association of CT and MRI utilisation with life expectancy at birth, three-year lag

| Modality | Model | n | k | $\beta$<br>(years) | 95% CI | p | p boot |
| --- | --- | --- | --- | --- | --- | --- | --- |
| CT | Country + year FE | 415 | 29 | 0.807 | 0.294 to<br>1.320 | 0.0020 | — |
| CT | + socioeconomic | 403 | 29 | 0.858 | 0.218 to 1.498 | 0.0086 | — |
| CT | **+ physicians**<br>**(principal)** | **339** | **29** | **0.849** | **0.162 to 1.536** | **0.0154** | **0.0340** |
| CT | + smoking (complete-case) | 105 | 29 | 1.664 | 0.700 to 2.627 | 0.0007 | 0.0115 |
| MRI | Country + year FE | 397 | 29 | 0.951 | -0.639 to 2.536 | 0.2414 | — |
| MRI | + socioeconomic | 385 | 29 | 0.826 | -0.697 to 2.348 | 0.2880 | — |
| MRI | + physicians | 324 | 28 | 0.231 | -1.141 to 1.602 | 0.7417 | — |
| MRI | + smoking (complete-case) | 101 | 28 | -0.193 | -2.258 to 1.872 | 0.8548 | — |

*Modality refers to CT or MRI examinations per 1,000 inhabitants, scaled per +100 examinations. "Socioeconomic" = log GDP per capita + log health expenditure per capita + population aged $\geq 65$ years. All models include country and year fixed effects with standard errors clustered by country. k = number of countries; p = cluster-robust; p boot = wild cluster bootstrap (Rademacher weights, null imposed, B = 1,999).*

### 3.4 Decomposition of the smoking-adjusted estimate

Adding smoking prevalence more than doubled the point estimate, from 0.85 to 1.66 years. Because smoking is reported for only 36% of country-years, this step changes the analytical sample as well as the adjustment set. Re-estimating the model **without** smoking in the identical 105-observation sample yielded β = 1.575 years (95% CI 0.635 to 2.515, p = 0.0010) (Table 3). Of the 0.82-year difference between the principal model and the smoking-adjusted model, 0.73 years (89%) is therefore attributable to change in sample composition and 0.09 years to the smoking adjustment itself. The smoking-adjusted estimate is accordingly reported as a sensitivity analysis in a restricted subsample, not as the study’s principal estimate.

**Table 3.** Decomposition of the difference between the principal and smoking-adjusted estimates

| Specification | n | $\beta$ (years) | 95% CI | p |
| --- | --- | --- | --- | --- |
| Full sample, model without smoking | 339 | 0.849 | 0.162 to 1.536 | 0.0154 |
| Restricted sample, model without smoking | 105 | 1.575 | 0.635 to 2.515 | 0.0010 |
| Restricted sample, model with smoking | 105 | 1.664 | 0.700 to 2.627 | 0.0007 |

### 3.5 Lead-exposure (placebo) analyses

CT utilisation three years in the future was associated with current life expectancy at β = 0.858 years (95% CI 0.151 to 1.565, p = 0.017), statistically indistinguishable from the corresponding three-year lag (β = 0.849, 95% CI 0.162 to 1.536, p = 0.015). The full lead–lag profile was essentially flat between lead 5 and lag 1 (Table 4, Figure 3). The association is therefore symmetric in time, and the lag structure carries no directional information — a result consistent with the smooth within-country evolution of the exposure documented in Section 3.2.

**Table 4.** Lead–lag profile: CT utilisation and life expectancy, extended adjusted mode

| Exposure timing | n | k | $\beta$ (years) | 95% CI | p |
| --- | --- | --- | --- | --- | --- |
| Lag 5 years (past) | 286 | 28 | 0.540 | 0.085 to 0.995 | 0.0200 |
| Lag 3 years (past) | 339 | 29 | 0.849 | 0.162 to 1.536 | 0.0154 |
| Lag 1 year (past) | 389 | 29 | 1.101 | 0.381 to 1.821 | 0.0027 |
| Same year | 415 | 29 | 0.920 | 0.242 to 1.598 | 0.0078 |
| Lead 1 year (future) | 435 | 29 | 0.803 | 0.175 to 1.431 | 0.0122 |
| Lead 3 years (future) | 449 | 29 | 0.858 | 0.151 to 1.565 | 0.0173 |
| Lead 5 years (future) | 436 | 29 | 0.802 | 0.283 to 1.322 | 0.0025 |

$k$ = number of countries. Lead models regress life expectancy in year $t$ on CT utilisation in year $t+j$ and are placebo tests: a causal effect cannot operate backwards in time.

**Figure 3.**
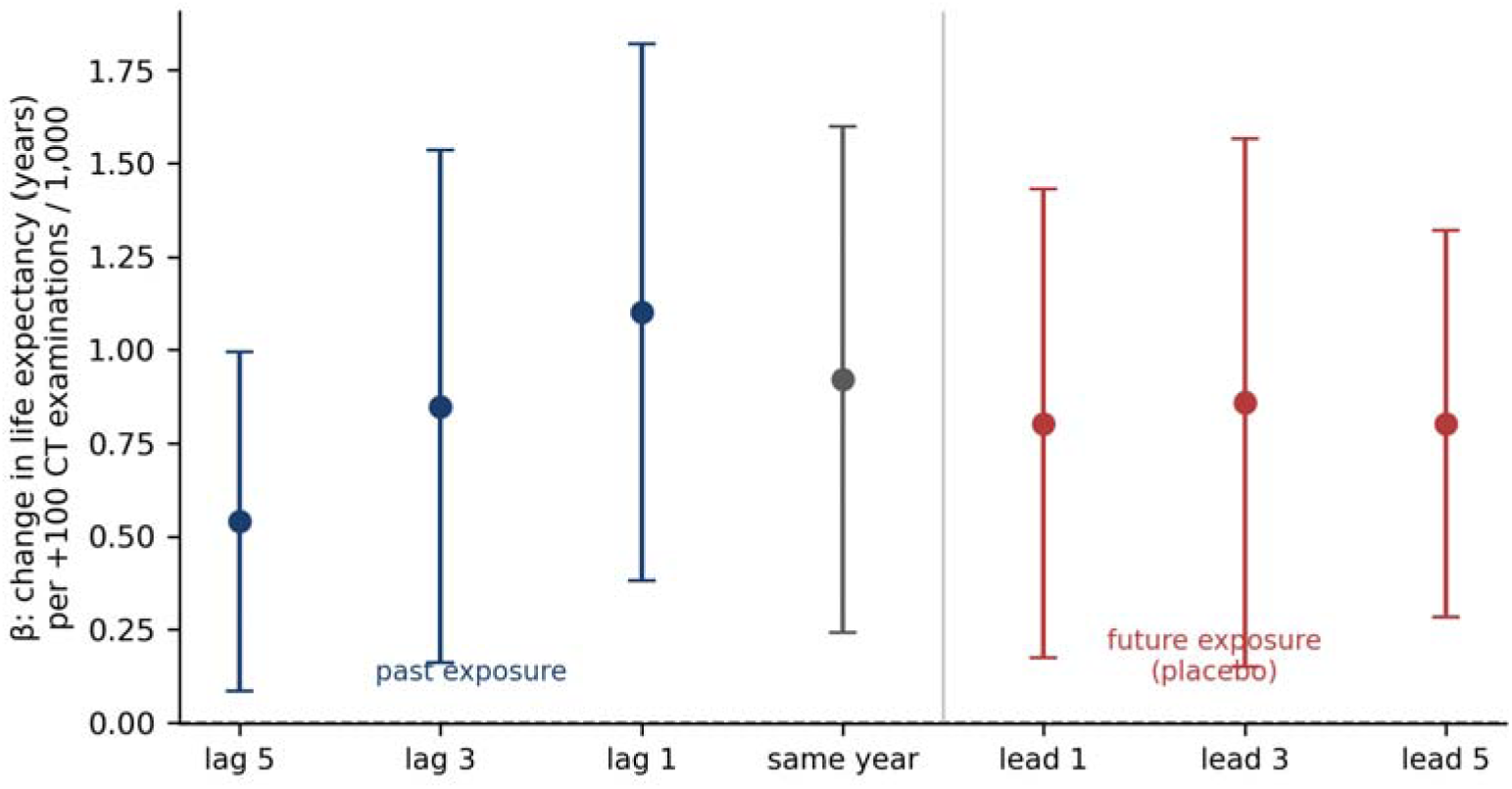
Lead–lag profile of the association between CT utilisation and life expectancy. Points are β coefficients with 95% confidence intervals from the extended adjusted fixed-effects model. Blue points use past exposure; red points use future exposure and constitute placebo tests. The profile is flat, indicating that the lag structure does not identify temporal direction.

### 3.6 Negative control exposures

Radiotherapy equipment density — a therapeutic technology with no diagnostic pathway to survival — was associated with life expectancy at β = 0.853 years per 10 additional units per million inhabitants (95% CI 0.321 to 1.384, p = 0.0017, n = 450), a coefficient of the same magnitude as, and estimated more precisely than, that for CT utilisation. PET scanner density (β = 2.520, 95% CI 0.212 to 4.828, p = 0.032) and log health expenditure per capita (β = 1.349, 95% CI 0.536 to 2.163, p = 0.0012) behaved similarly. In contrast, physician density showed no association whatsoever (β = 0.033, 95% CI −0.135 to 0.201, p = 0.700), as did CT scanner density and mammograph density (Table 5, Figure 4).

**Table 5.** Negative control exposures and life expectancy at birth, three-year lag, extended adjusted model

| Exposure (scaling) | Type | n | $\beta$<br>(years) | 95% CI | p |
| --- | --- | --- | --- | --- | --- |
| CT examinations / 1,000 (+100) | Diagnostic imaging | 339 | 0.849 | 0.162 to 1.536 | 0.0154 |
| MRI examinations / 1,000 (+100) | Diagnostic imaging | 324 | 0.231 | $-1.141$ to $1.602$ | 0.7417 |
| PET examinations / 1,000 (+100) | Diagnostic imaging | 259 | $-0.266$ | $-14.975$ to $14.442$ | 0.9717 |
| CT scanners / million (+10) | Diagnostic imaging | 565 | 0.258 | $-0.084$ to $0.601$ | 0.1395 |
| MRI units / million (+10) | Diagnostic imaging | 532 | 0.774 | 0.002 to 1.546 | 0.0496 |
| Radiotherapy equipment / million (+10) | Negative control | 450 | 0.853 | 0.321 to 1.384 | 0.0017 |
| Mammographs / million (+10) | Negative control | 435 | 0.233 | $-0.073$ to $0.539$ | 0.1349 |
| Gamma cameras / million (+10) | Negative control | 439 | 0.649 | $-0.074$ to $1.372$ | 0.0787 |
| PET scanners / million (+10) | Negative control | 468 | 2.520 | 0.212 to 4.828 | 0.0324 |
| Physicians / 1,000 (+1) | Negative control | 777 | 0.033 | $-0.135$ to $0.201$ | 0.7004 |
| log health expenditure per capita (+1) | Negative control | 729 | 1.349 | 0.536 to 2.163 | 0.0012 |

*Negative control exposures were selected as national health-system resources that share the determinants of imaging investment but have no diagnostic-imaging pathway to population survival. Models for physician density and health expenditure exclude that variable from the covariate set.*

**Figure 4.**
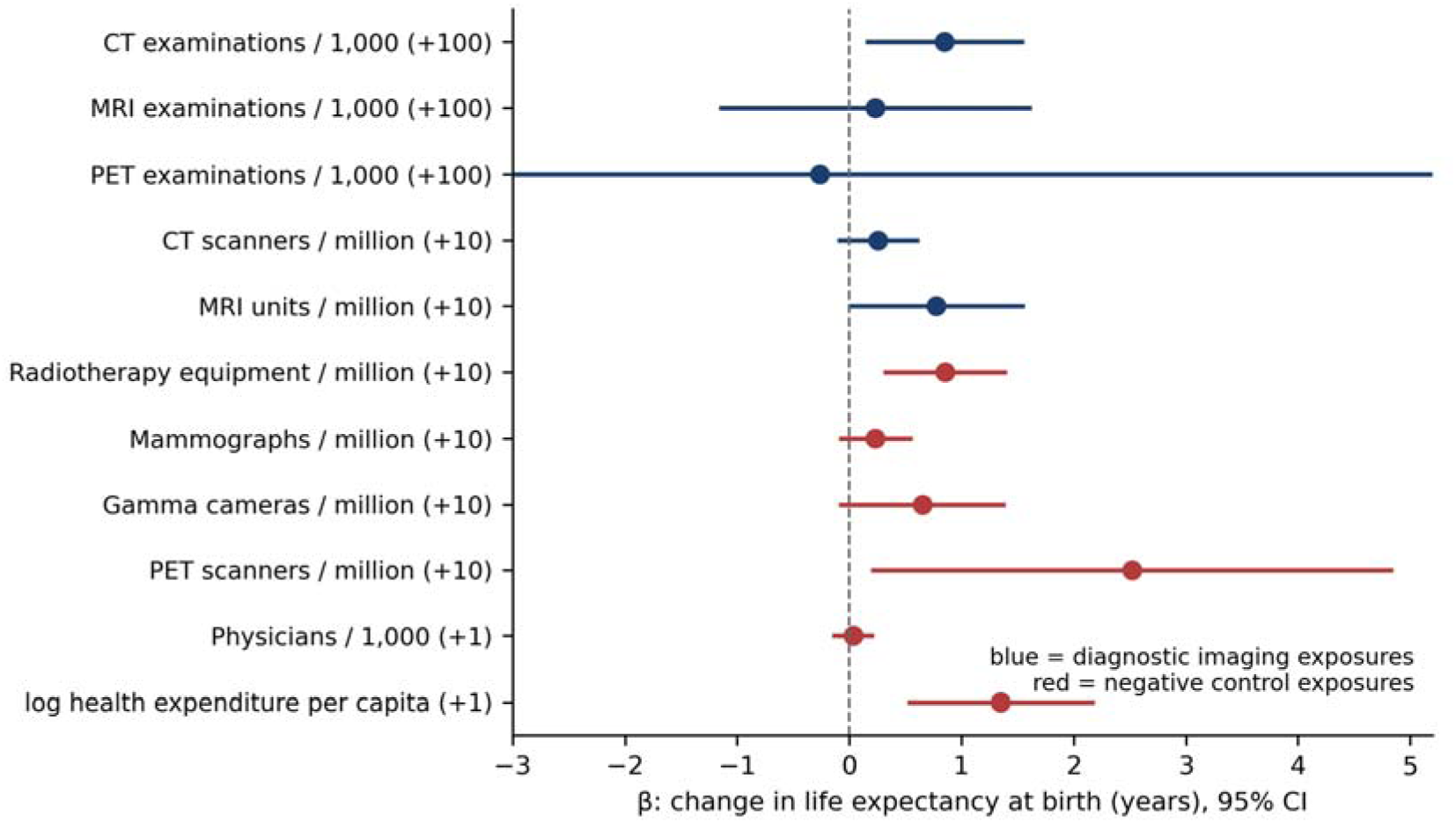
Negative control exposures and life expectancy at birth. Points are β coefficients with 95% confidence intervals from the extended adjusted fixed-effects model at a three-year lag. Blue: diagnostic imaging exposures. Red: negative control exposures. The confidence interval for PET examinations extends beyond the plotted range.

### 3.7 Treatable and preventable mortality

CT utilisation was substantially more strongly associated with preventable than with treatable mortality. In the extended adjusted model at a three-year lag, an increase of 100 CT examinations per 1,000 inhabitants was associated with β = −43.28 deaths per 100,000 for preventable mortality (95% CI −66.36 to −20.20, p = 0.0002) but only β = −13.49 for treatable mortality (95% CI −33.70 to 6.73, p = 0.191) (Table 6). A similar asymmetry was observed for MRI utilisation.

**Table 6.**
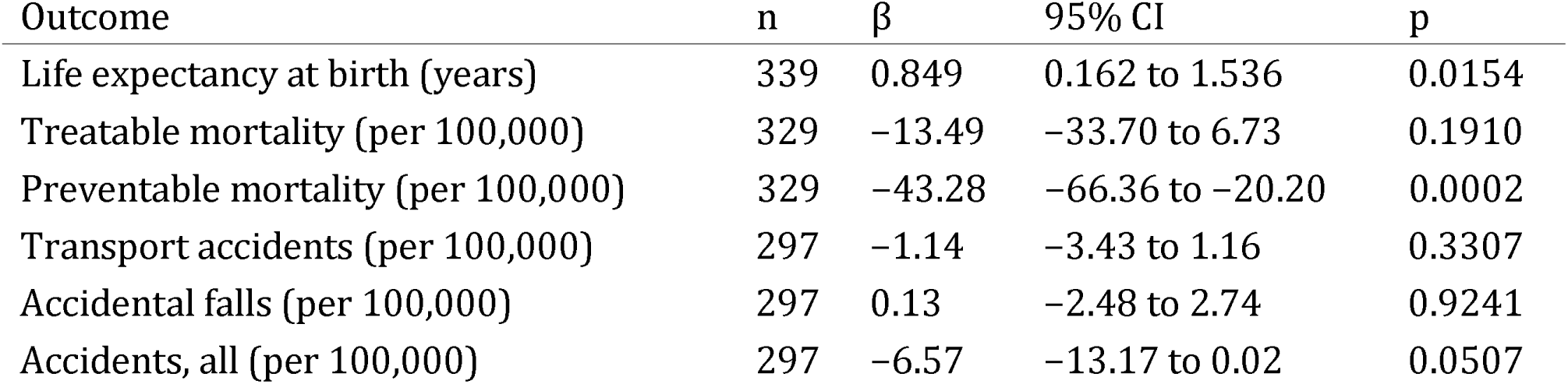
CT utilisation (three-year lag) and mortality outcomes, extended adjusted model

| Outcome | n | $\beta$ | 95% CI | p |
| --- | --- | --- | --- | --- |
| Life expectancy at birth (years) | 339 | 0.849 | 0.162 to 1.536 | 0.0154 |
| Treatable mortality (per 100,000) | 329 | -13.49 | -33.70 to 6.73 | 0.1910 |
| Preventable mortality (per 100,000) | 329 | -43.28 | -66.36 to -20.20 | 0.0002 |
| Transport accidents (per 100,000) | 297 | -1.14 | -3.43 to 1.16 | 0.3307 |
| Accidental falls (per 100,000) | 297 | 0.13 | -2.48 to 2.74 | 0.9241 |
| Accidents, all (per 100,000) | 297 | -6.57 | -13.17 to 0.02 | 0.0507 |

### 3.8 Cause-specific and negative control outcomes

Cause-specific data were available for all 38 countries with approximately 85% completeness. Several nominally significant associations emerged before multiplicity correction, including CT utilisation and lung cancer mortality at lags 0, 1 and 3 years (lag 3: β = −3.52 per 100,000, 95% CI −6.48 to −0.56, p = 0.0196), CT scanner availability and ischaemic heart disease mortality, MRI availability and transport-accident mortality, and CT utilisation and accident mortality at lag 5. The exploratory family comprised 112 comparisons; after Benjamini–Hochberg correction no inverse imaging–mortality association remained significant at q < 0.05.

Outcomes with no plausible diagnostic-imaging pathway behaved as negative controls would be expected to if the survival association were modality-specific — but so, largely, did the imaging-sensitive outcomes. CT utilisation was unassociated with transport-accident mortality (β = −1.14, 95% CI −3.43 to 1.16, p = 0.331) and accidental falls (β = 0.13, 95% CI −2.48 to 2.74, p = 0.924), and only borderline for accidents overall (β = −6.57, 95% CI −13.17 to 0.02, p = 0.051) (Table 6).

### 3.9 Restricted secondary analysis

Six biologically motivated associations were evaluated with the full adjustment set including smoking, with Holm correction (Table 7). Only CT utilisation and life expectancy at a three-year lag reached significance under cluster-robust inference (β = 1.664, 95% CI 0.700 to 2.627, p = 0.00071; Holm-adjusted p = 0.0043). Under wild cluster bootstrap inference the unadjusted p for this model was 0.0115, which corresponds to a Holm-adjusted p of approximately 0.069 across the six comparisons. The restricted analysis therefore does not provide inference that is robust to both multiplicity and small-cluster correction. No cause-specific hypothesis was supported.

**Table 7.** Smoking-adjusted restricted hypothesis-driven validation analyses

| H | Exposure | Outcome | Lag | n | $\beta$ | 95% CI | p | Holm p |
| --- | --- | --- | --- | --- | --- | --- | --- | --- |
| H1 | CT exams | Lung cancer | 3 | 72 | -1.994 | -9.803 to 5.814 | 0.6167 | 1.0000 |
| H2 | CT exams | Cerebrovascular | 1 | 86 | -3.821 | -43.696 to 36.053 | 0.8510 | 1.0000 |
| H3 | MRI exams | Cerebrovascular | 1 | 83 | 26.104 | -33.062 to 85.271 | 0.3872 | 1.0000 |
| H4 | CT exams | Accidents | 1 | 84 | -15.073 | -33.727 to 3.580 | 0.1132 | 0.5662 |
| H5 | CT exams | Life expectancy | 3 | 105 | 1.664 | 0.700 to 2.627 | 0.0007 | 0.0043 |
| H6 | MRI exams | Life expectancy | 3 | 101 | -0.193 | -2.258 to 1.872 | 0.8548 | 1.0000 |

*Hypotheses were selected after exploratory analyses and are not independently pre-registered confirmatory tests. Under wild cluster bootstrap inference, the Holm-adjusted $p$ for H5 is approximately 0.069.*

### 3.10 First-difference and country-trend models

In first-difference models the annual change in CT utilisation was not associated with the annual change in life expectancy (β = 0.14 years, p = 0.627); MRI utilisation showed a larger but non-significant association (β = 0.74, p = 0.234). For treatable mortality the strongest first-difference signal was for MRI utilisation (β = −7.43 per 100,000, p = 0.058).

When country-specific linear time trends were added to the two-way fixed-effects specification, all principal associations were attenuated to the null, and — critically — so were the negative control associations: CT utilisation β = 0.291 (95% CI −0.114 to 0.696, p = 0.159), radiotherapy equipment β = −0.063 (95% CI −0.776 to 0.651, p = 0.864), MRI utilisation β = −0.153 (p = 0.789), PET scanners β = −0.088 (p = 0.936). Imaging and non-imaging capacity measures therefore behave identically under trend adjustment, as would be expected if all are proxies for a single slowly evolving national process (Table 8, Figure 5).

**Table 8.** Country-specific linear trend models, three-year lag, life expectancy

| Exposure | n | $\beta$ (years) | 95% CI | p |
| --- | --- | --- | --- | --- |
| CT examinations / 1,000 (+100) | 339 | 0.291 | -0.114 to 0.696 | 0.159 |
| MRI examinations / 1,000 (+100) | 324 | -0.153 | -1.276 to 0.969 | 0.789 |
| Radiotherapy equipment / million (+10) | 450 | -0.063 | -0.776 to 0.651 | 0.864 |
| PET scanners / million (+10) | 468 | -0.088 | -2.224 to 2.049 | 0.936 |

**Figure 5.**
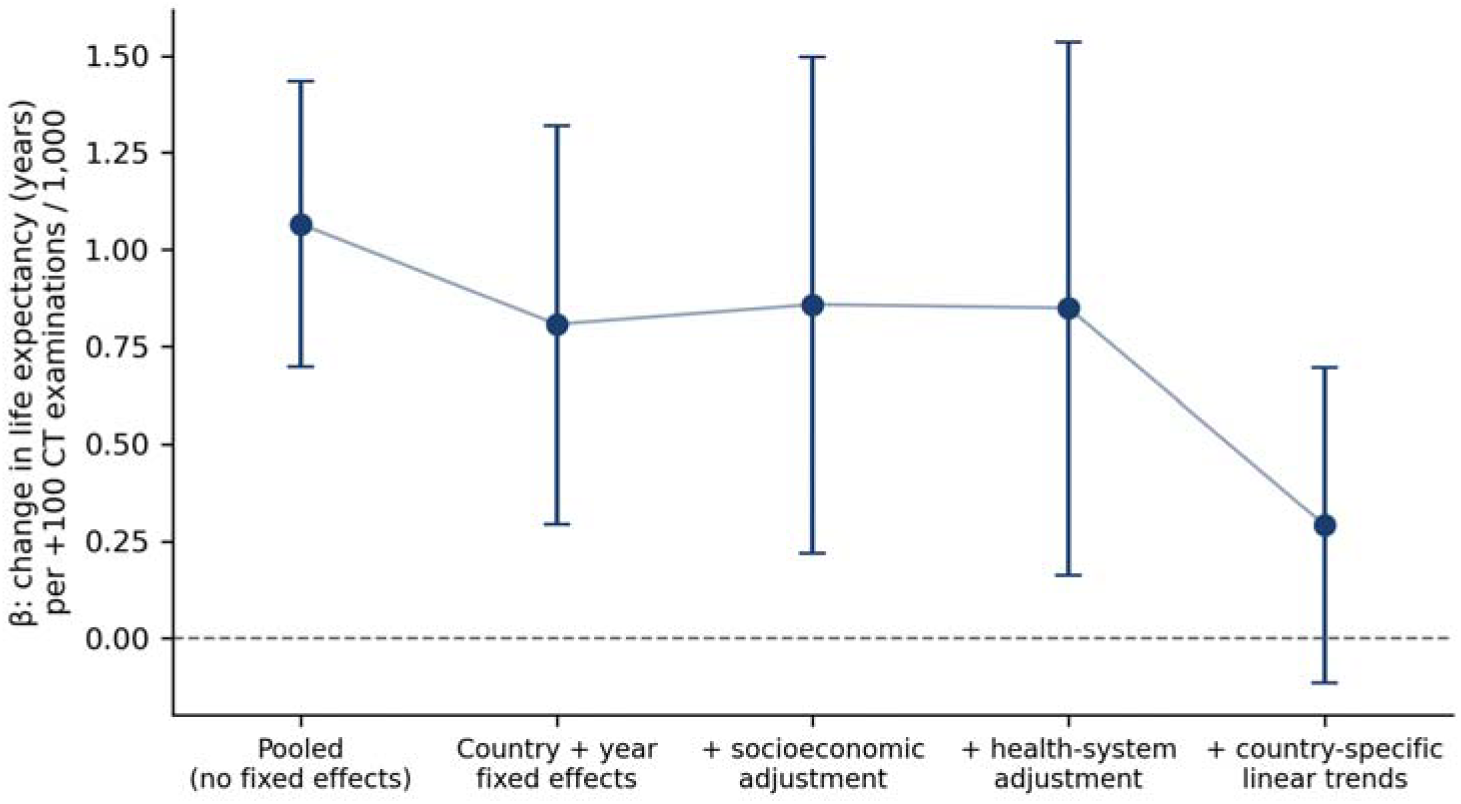
Progressive attenuation of the CT utilisation–life expectancy association. Points are β coefficients with 95% confidence intervals for +100 CT examinations per 1,000 inhabitants at a three-year lag, under successively stronger control for confounding.

### 3.11 Summary of findings

Four patterns emerged. First, imaging utilisation showed stronger associations with population outcomes than scanner availability. Second, the association between CT utilisation and life expectancy was stable across sequential socioeconomic and health-system adjustment. Third, every falsification test designed to isolate a modality-specific effect failed: the association was symmetric in lead and lag, was reproduced in equal magnitude by a therapeutic technology, was concentrated in preventable rather than treatable mortality, and did not survive country-specific trend adjustment. Fourth, cause-specific associations did not survive multiplicity correction.

## 4. Discussion

In this longitudinal analysis of 38 OECD countries over 25 years, national CT utilisation was a robust ecological correlate of life expectancy, whereas scanner availability was not. The central contribution of this study, however, is not the correlation itself — which has been reported before in other settings [1–3] — but the demonstration that it does not behave as a causal relationship under falsification.

### 4.1 Imaging utilisation versus imaging capacity

The clearest robust pattern was the divergence between technology availability and technology use. CT and MRI units per million inhabitants were only weakly associated with survival after fixed-effects adjustment, whereas examination volumes retained stronger associations. This is clinically plausible: the presence of equipment does not imply timely access, appropriate referral pathways, sufficient staffing, workflow integration or effective downstream treatment. From a health-services perspective, radiological infrastructure alone may carry limited population-level meaning unless translated into delivered service; the routine reporting of capacity and volume as separate national indicators [33] is therefore not redundant, and our results suggest the two carry different information. This finding is robust to all analyses reported here and is, in our view, the most directly actionable result of the study.

### 4.2 Why the survival association is unlikely to be modality-specific

Three independent lines of evidence converge.

First, the lead–lag profile is flat. CT utilisation three and five years *future* in the future predicted current life expectancy as well as past utilisation. A causal effect cannot operate backwards in time, so the lagged estimate cannot be interpreted as evidence of temporal precedence [24]. The mechanism is transparent: within-country CT utilisation is close to a smooth trend (autocorrelation 0.64 at three years), so lagged and led versions of the exposure are nearly the same variable, and the design has essentially no power to distinguish them. This also implies that the choice of a three-year lag in our own and in prior analyses is a presentational convention, not an identification strategy.

Second, negative control exposures reproduce the finding. Under the standard logic of negative controls, an exposure that shares the confounding structure of the exposure of interest but lacks its causal pathway should be null if the observed association is causal and non-null if it is confounded [17,18]. Radiotherapy equipment density — capital-intensive, technologically advanced, and entirely therapeutic — showed an association of the same magnitude and greater precision than CT utilisation. PET scanner density and health expenditure per capita behaved likewise. Importantly, the pattern is not indiscriminate: physician density showed no association at all, and neither did CT scanner or mammograph density. The proxy being measured therefore appears to be specifically the accumulation of capital-intensive technological capacity within a health system, rather than health-system size or clinical workforce in general. Under trend adjustment, imaging and non-imaging capacity measures were attenuated in parallel, which is the signature of several indicators tracking a single slow process rather than several distinct causal channels.

Third, the outcome pattern points the wrong way. If imaging acted primarily through diagnosis and treatment, the strongest association should be with *treatable* mortality — deaths avoidable through timely and effective healthcare. Instead the association was three times larger and far more precise for *preventable* mortality, a category defined in the OECD/Eurostat classification as avoidable through public health and primary prevention rather than through healthcare delivery [11], and dominated by tobacco, alcohol and road traffic, in which diagnostic imaging has no plausible mechanism. Both avoidable-mortality outcomes are, however, restricted to deaths below 75 years, which the contrast does not fully control for. Cause-specific analyses in imaging-decisive conditions did not survive multiplicity correction.

### 4.3 Imaging utilisation as a health-system marker

Taken together, these results support an interpretation in which national CT utilisation functions as a high-resolution marker of health-system capital development. Countries that expand and integrate CT services concurrently improve emergency care, specialist access, hospital throughput, referral coordination and oncology pathways, and it is this composite that tracks longevity. Imaging utilisation is an unusually good marker of that composite because it is measured annually, comparably and with fine gradation — better, in this dataset, than physician density or scanner counts.

This is a weaker claim than “imaging extends life,” but it is a more defensible one, and it has its own policy value. A national imaging utilisation series can serve as a monitorable indicator of diagnostic access and system throughput, without requiring the assumption that increasing the number of examinations would itself produce survival gains. Indeed, our results provide no support for that assumption, and the well-documented harms of imaging expansion — radiation exposure, incidental findings, overdiagnosis, downstream invasive procedures and cost [28–30] — argue strongly against inferring it.

### 4.4 Why MRI diverged from CT

MRI utilisation showed no consistent relationship with life expectancy. CT is more broadly used in acute and life-threatening conditions, whereas MRI is concentrated in neurological, musculoskeletal, oncological and elective pathways whose mortality impact is more disease-specific and slower; utilisation data from large health systems show the two modalities growing at different rates and serving different case mixes [28]. MRI throughput is also more constrained by examination time and staffing, making examination counts less comparable across systems. These explanations are plausible but are not tested by our data and should be regarded as hypotheses rather than findings. Under the marker interpretation, CT utilisation may simply be a better-measured and more responsive index of overall system throughput than MRI utilisation. The divergence should not be read as evidence that MRI has less clinical value.

### 4.5 Relation to previous work

Our findings are consistent in sign with previous state-level analyses reporting faster longevity growth where advanced imaging use grew fastest [1–3], but they qualify their interpretation substantially. Those analyses did not report lead-exposure placebo models or negative control exposures, and the methodological objections raised against them at the time [5] were largely about specification rather than about falsification. When we apply both to a comparable design, the survival association survives adjustment but fails falsification. We would suggest that reported estimates of the years of life attributable to imaging in aggregate data — including our own — should be treated as upper bounds on a composite health-system effect rather than as modality-specific quantities.

### 4.6 Implications for evaluation of artificial intelligence in radiology

Much current evaluation of AI in radiology relies on intermediate endpoints: diagnostic accuracy, reading time, prioritisation, workflow efficiency, and the reporting quality of such studies has itself been questioned [31]. Randomised evaluation against clinically meaningful endpoints is feasible and has begun in screening [32]. Our results suggest a caution for any attempt to bridge from such endpoints to population outcomes using ecological data. If national imaging utilisation — a far more direct measure of imaging activity than AI deployment — cannot be shown to act on survival independently of general health-system development, then aggregate analyses of AI adoption and population outcomes will be still less identifiable. Where imaging has been shown to change mortality, it has been through randomised designs with defined populations and endpoints [25–27], and the same standard should apply to AI. Demonstrating population benefit from AI-enabled imaging will require patient-level linkage or natural-experiment designs, not cross-national correlation.

## 5. Limitations

The analysis is ecological and based on aggregated national data; associations cannot be extrapolated to individual patients, and the study is subject to the well-described biases of ecological inference [22,23].

Treatable and preventable mortality are restricted by definition to deaths below 75 years, whereas life expectancy at birth reflects mortality at all ages. The contrast between these outcomes is therefore not only a contrast of causal pathway but partly of age range, and should be interpreted accordingly.

Residual confounding remains likely despite country and year fixed effects and adjustment for GDP, health expenditure, physician density, population ageing and smoking. Unmeasured determinants include educational attainment, income inequality, insurance coverage, hospital quality, primary-care access, emergency-service organisation, screening intensity, treatment availability, alcohol consumption and physical activity.

Reverse causality cannot be excluded, and the lead-exposure analyses reported here indicate that lagging the exposure does not resolve it.

Imaging utilisation data were substantially less complete than outcome data: CT and MRI examination rates were available for approximately half of possible country-years across 29 countries, so utilisation analyses rest on an unbalanced panel that may be selectively reported. Smoking prevalence was available for only 36% of country-years, and we have shown that smoking-adjusted estimates are dominated by the resulting change in sample composition.

The exposure is continuous and non-staggered, so the two-way fixed-effects estimator imposes linearity and effect homogeneity; under heterogeneous effects the estimand is a weighted average that need not correspond to any policy-relevant contrast [14–16].

The number of clusters is modest (29 countries), and cluster-robust inference is correspondingly fragile; we report wild cluster bootstrap p values for the principal models, which are approximately two- to sixteen-fold larger than the cluster-robust values.

National imaging counts carry no information on clinical indication, appropriateness, urgency, diagnostic yield, disease stage or downstream treatment, and do not distinguish inpatient from outpatient, emergency from elective, or diagnostic from follow-up examinations. National differences in coding, reporting practice, equipment definitions and provider coverage introduce measurement heterogeneity.

Negative control exposures are informative but imperfect: radiotherapy equipment, PET scanners and gamma cameras are not entirely independent of diagnostic pathways, and a shared association could in principle arise from a common causal channel rather than from confounding. Their value lies in the absence of a *diagnostic-imaging-specific* mechanism, not in the absence of any mechanism.

Country-specific trend models may absorb genuine slow-acting effects along with confounding, and their null results should not be read as proof of no effect.

Finally, the study does not quantify the harms or opportunity costs of imaging, and its findings must not be read as support for increasing imaging volume without regard to appropriateness.

## 6. Conclusions

Across 38 OECD countries between 2000 and 2024, higher national CT utilisation was associated with greater life expectancy after adjustment for socioeconomic development, healthcare expenditure, physician density, population ageing and smoking, whereas scanner availability alone was not. This distinction between technology availability and technology use is robust and policy-relevant: infrastructure without delivered service carries little population-level signal.

The survival association itself, however, does not withstand falsification. It is symmetric in lead and lag, is reproduced in equal magnitude by a purely therapeutic technology, is concentrated in mortality categories in which imaging has no mechanism, and disappears under country-specific trend adjustment. National CT utilisation is best understood as a marker of health-system capital development rather than an independent determinant of longevity.

Population-level ecological data of this kind can describe how imaging tracks health-system performance. It cannot establish that imaging extends life, and claims to that effect should not be based on it. Determining whether expanded imaging access itself contributes to survival will require patient-level linkage and natural-experiment designs.

## Supporting information

strobe table

## Data Availability

article code and data are avalable at zenodo: https://doi.org/10.5281/zenodo.22257963

## 7. Declarations

Ethics. This study analysed exclusively publicly available, aggregated, country-level statistics published by the OECD and the World Bank. No individual-level, patient-level or otherwise identifiable data were accessed or analysed, no human participants were involved, and no biological material was used. The study therefore does not constitute research on human subjects under applicable national and institutional regulations, and formal review by a research ethics committee was not required; informed consent was not applicable.

Data availability. All source data are publicly available from OECD Health Statistics and the World Bank World Development Indicators. The constructed country-year panel, the panel-construction script and the complete analysis and figure code are openly deposited at [Zenodo DOI: **10.5281/zenodo.22257963**. A completed STROBE checklist is provided as a supplementary file].

## Reporting guideline

STROBE (cross-sectional/ecological adaptation) [21].

