## Supplementary material for "Marker or mechanism? Medical imaging utilisation and population life expectancy in 38 OECD countries, 2000–2024 The RAD-LIFE study": strobe table

### STROBE checklist — RAD-LIFE

**Design:** longitudinal ecological panel study (country-year unit of analysis). The STROBE cross-sectional checklist is used, with adaptations noted where items refer to individual participants. Page numbers refer to the preprint PDF, version 1.0.

**Status key:** *Yes* = addressed; *Partial* = partially addressed, see note; *n/a* = not applicable to an ecological design.

#### Title and abstract

| Item | STROBE recommendation | Status | Where | Note |
| --- | --- | --- | --- | --- |
| 1a | Indicate the study design in the title or abstract | Yes | Title, p. 1; Abstract Methods, p. 1 | “longitudinal analysis”; Abstract states “longitudinal ecological panel study” |
| 1b | Provide in the abstract an informative and balanced summary | Yes | p. 1 | Includes the negative falsification results and the deflationary conclusion |

#### Introduction

| Item | STROBE recommendation | Status | Where | Note |
| --- | --- | --- | --- | --- |
| 2 | Explain the scientific background and rationale | Yes | §1, p. 2 | Includes prior claims [1–4] and the existing methodological critique [5] |
| 3 | State specific objectives, including any prespecified hypotheses | Partial | §1, p. 2; §2.14, p. 6 | Objectives and falsification strategies stated. The restricted analyses were selected after inspection of exploratory results and are explicitly labelled as such, not as prespecified confirmatory tests |

#### Methods

| Item | STROBE recommendation | Status | Where | Note |
| --- | --- | --- | --- | --- |
| 4 | Present key elements of study design early in the paper | Yes | §2.1, p. 3 |  |
| 5 | Describe the setting, locations and relevant dates | Yes | §2.1, p. 3 | 38 OECD countries, 2000–2024, 950 possible country-years |
| 6 | Eligibility criteria, sources and methods of selection of participants | Partial | §2.1, p. 3; §2.6, p. 4 | Adapted: the units are countries, not participants. All 38 OECD members were eligible; inclusion in any given model was determined by simultaneous availability of the required variables (unbalanced complete-case panel) |
| 7 | Clearly define all outcomes, exposures, predictors, potential confounders and effect modifiers | Yes | §2.3–2.5, pp. 4 | Avoidable-mortality definitions follow the OECD/Eurostat joint lists and are restricted to deaths below 75 years |
| 8 | For each variable, give sources of data and details of methods of assessment | Yes | §2.2, p. 3; Table 1, p. 7 | OECD Health Statistics and World Bank WDI; Table 1 gives per-variable definitions and completeness |
| 9 | Describe any efforts to address potential sources of bias | Yes | §2.7, p. 5; §2.9–2.12, pp. 5–6 | Country and year fixed effects; lead-exposure placebo models; negative control exposures and outcomes; country-specific trend models; first-difference models |
| 10 | Explain how the study size was arrived at | Partial | §2.1, p. 3; §3.1, p. 7 | No a priori sample size calculation: the panel is defined by the population of OECD countries and by data availability. Analytic n per model is reported in every results table |
| 11 | Explain how quantitative variables were handled and why | Yes | §2.3, p. 4; §2.5, p. 4 | Exposures kept continuous and scaled for interpretability (+100 examinations/1,000; +10 units/million); GDP and health expenditure log-transformed with rationale |
| 12a | Describe all statistical methods, including those used to control for confounding | Yes | §2.7–2.8, p. 5 | Two-way fixed effects with sequential adjustment; limits of TWFE under continuous heterogeneous exposure stated explicitly |
| 12b | Describe any methods used to examine subgroups and interactions | Partial | §2.11, p. 6; §2.14, p. 6 | No formal interaction testing. Cause-specific analyses and the restricted subsample serve this role and are labelled exploratory |
| 12c | Explain how missing data were addressed | Yes | §2.6, p. 4; §3.4, p. 10 | Complete-case at model level, no imputation; the effect of complete-case restriction on the smoking-adjusted estimate is quantified in §3.4 |
| 12d | If applicable, describe analytical methods taking account of sampling strategy | n/a | — | No sampling: the study covers the full OECD membership |
| 12e | Describe any sensitivity analyses | Yes | §2.9–2.12, pp. 5–6; §2.15, p. 6 | Lags 0/1/3/5 and leads 1/3/5; negative controls; country-specific trends; first differences; wild cluster bootstrap |

#### Results

| Item | STROBE recommendation | Status | Where | Note |
| --- | --- | --- | --- | --- |
| 13a | Report numbers of individuals at each stage of the study | Partial | §3.1, p. 7; Table 1, p. 7 | Adapted: country-years observed against country-years possible, per variable |
| 13b | Give reasons for non-participation at each stage | Partial | §3.1, p. 7; §5, p. 18 | Missingness reflects national reporting practice to the OECD; not attributable at country level, and named as a limitation (potential selective reporting) |
| 13c | Consider use of a flow diagram | Partial | Table 1, p. 7 | No flow diagram; Table 1 serves this function. A CONSORT-style flow figure could be added if the target journal requests one |
| 14a | Give characteristics of study participants and information on exposures and potential confounders | Yes | §3.1–3.2, pp. 7–8; Figure 1, p. 8 |  |
| 14b | Indicate the number of participants with missing data for each variable of interest | Yes | Table 1, p. 7 | Per-variable completeness for all 16 variables |
| 15 | Report numbers of outcome events or summary measures | Yes | §3.1–3.2, pp. 7–8 | Outcomes are national rates and life expectancy; distributions and trends reported |
| 16a | Give unadjusted and confounder-adjusted estimates and their precision; make clear which confounders were adjusted for and why | Yes | §3.3, p. 9; Table 2, p. 9; Figure 5, p. 15 | Pooled, fixed-effects-only and sequentially adjusted estimates all reported with 95% CI |
| 16b | Report category boundaries when continuous variables were categorized | n/a | — | No categorisation; all exposures analysed continuously |
| 16c | If relevant, consider translating estimates of relative risk into absolute risk | Yes | Tables 2–8 | Estimates are already on an absolute scale (years of life expectancy; deaths per 100,000) |
| 17 | Report other analyses done — subgroups, interactions, sensitivity analyses | Yes | §3.4–3.10, pp. 10–15; Tables 3–8 |  |

#### Discussion

| Item | STROBE recommendation | Status | Where | Note |
| --- | --- | --- | --- | --- |
| 18 | Summarise key results with reference to study objectives | Yes | §3.11, p. 15; §4 opening, p. 16 |  |
| 19 | Discuss limitations, taking into account sources of potential bias or imprecision, and the direction and magnitude of any potential bias | Yes | §5, p. 18 | Ecological inference, residual confounding, reverse causality, selective reporting, small number of clusters, TWFE under heterogeneity, age restriction of avoidable mortality, limits of the negative controls themselves |
| 20 | Give a cautious overall interpretation considering objectives, limitations, multiplicity of analyses, results from similar studies and other relevant evidence | Yes | §4.2–4.5, pp. 16–18 | Multiplicity handled by Benjamini–Hochberg and Holm; relation to prior work and to its published critique addressed in §4.5 |
| 21 | Discuss the generalisability (external validity) of the study results | Partial | §5, p. 18; §4.3, p. 17 | Restricted to OECD countries; heterogeneous national reporting standards noted. Generalisability beyond OECD settings is not addressed in a dedicated passage and could be strengthened |

#### Other information

| Item | STROBE recommendation | Status | Where | Note |
| --- | --- | --- | --- | --- |
| 22 | Give the source of funding and the role of the funders for the present study and, if applicable, for the original study on which the present article is based | Partial | §7, p. 20 | **Incomplete — must be finalised before posting or submission.** Funding and competing-interest statements are placeholders. Given the documented industry involvement in this literature, both must be explicit |
